# Public health relevant consequences of the COVID-19 pandemic on malaria in sub-Saharan Africa: A scoping review

**DOI:** 10.1101/2021.06.17.21258914

**Authors:** Anna-Katharina Heuschen, Guangyu Lu, Oliver Razum, Alhassan Abdul-Mumin, Osman Sankoh, Lorenz von Seidlein, Umberto D’Alessandro, Olaf Müller

**Affiliations:** Institute of Global Health, Medical School, Ruprecht-Karls-University Heidelberg, Germany; Department of Public Health, Medical College, Yangzhou University, China; Department of Epidemiology and International Public Health, School of Public Health, Bielefeld University, Germany; University for Development Studies, School of Medicine, Department of Paediatrics and Child Health, Tamale, Ghana; Statistics Sierra Leone, Tower Hill, Freetown, Sierra Leone; School of Public Health, Faculty of Health Sciences, University of the Witwatersrand, Johannesburg, South Africa; Mahidol Oxford Tropical Medicine Research Unit, Faculty of Tropical Medicine, Mahidol University, Bangkok, Thailand; MRC The Gambia

**Keywords:** COVID-19, coronavirus, malaria, pandemic, sub-Saharan Africa, public health, global health

## Abstract

**Background:** The COVID-19 pandemic has resulted in unprecedented challenges to health systems worldwide, including the control of non-COVID-19 diseases. Malaria cases and deaths may increase due to the direct and indirect effects of the pandemic in malaria endemic countries, particularly in sub-Saharan Africa (SSA).

**Objectives:** This scoping review aims to summarize information on public health relevant effects of the COVID-19 pandemic on the malaria situation in SSA.

**Methods:** Review of publications and manuscripts on preprint servers, in peer-reviewed journals and in grey literature documents from December 1, 2019, to June 9, 2021. A structured search was conducted on different databases using predefined eligibility criteria for the selection of articles.

**Results:** A total of 51 papers have been included in the analysis. Modeling papers have predicted a significant increase in malaria cases and malaria deaths in SSA due to the effects of the COVID-19 pandemic. Many papers provided potential explanations for expected COVID-19 effects on the malaria burden; these ranged from relevant diagnostical and clinical aspects, to reduced access to health care services, impaired availability of curative and preventive commodities and medications, and effects on malaria prevention campaigns. Compared to previous years, fewer country reports provided data on the actual number of malaria cases and deaths in 2020, with mixed results. While highly endemic countries reported evidence of decreased malaria cases in health facilities, low endemic countries reported overall higher numbers of malaria cases and deaths in 2020.

**Conclusions:** The findings from this review provide evidence for a significant but diverse impact of the COVID-19 pandemic on malaria in SSA. There is the need to further investigate the public health consequences of the COVID-19 pandemic on the malaria burden.

## Introduction

The emergence of SARS-CoV-2 in China by the end of 2019 has led to the largest pandemic in recent human history (1, 2). By June 14, 2021, there were some 176 million confirmed cases of COVID-19, including 3.8 million deaths, reported to the World Health Organization (WHO) (3). The COVID-19 epidemic waves show variable dynamics in the different WHO Regions, with the highest burden in the American, European and South-East Asian Regions (3, 4). The latter has recently shown particularly high incidence rates, and India is now reporting the second highest number of confirmed cases after the USA (3). In contrast, the African and the Western Pacific WHO Regions continue to report only relatively low numbers of cases and deaths (3, 4).

It was initially predicted that Africa would be the worst affected region by the COVID-19 pandemic because of its weak health systems, prevailing poverty, and the high burden of other infectious diseases (5, 6). However, by the end of 2020, only about 3.5% of the global number of COVID-19 cases and deaths were reported from this continent, which is home to 17% of the world’s population (3, 7). Overall, the epidemiology of COVID-19 in Africa remains puzzling (5). By June 14, 2021, there were some 3.6 million COVID-19 cases and 89,000 deaths reported from the entire continent, and most of these were from its northern and southern regions (8, 9). Potential explanations for such a situation are incomplete data due to much lower testing capacities, a significantly younger population, overall lower population mobility, cross-reactive immunity or immunomodulation due to high prevalence of other infectious agents, and effects of public health responses (5, 7, 10). First findings from SARS-CoV-2 seroprevalence surveys support the evidence for significant under-reporting and for a predominance of asymptomatic and mild cases (11, 12). Nevertheless, it appears that the second epidemic wave has hit the African continent more severely than the first one, possibly explained by the emergence of more transmissible SARS-CoV-2 variants (7, 13).

Globally, malaria is still the most important parasitic disease and responsible for a quarter of all deaths among Under Five children in sub-Saharan Africa (SSA) (14, 15). The efforts for global malaria control and elimination have achieved large successes during the last two decades, but progress has stalled in recent years, and the COVID-19 pandemic could largely reverse the overall trend (16, 17). This review aims to summarize currently available data and understanding of the direct and indirect effects of the COVID-19 pandemic on the malaria burden in SSA.

## Methods

### Search strategy and selection criteria

Due to the complex topic and the different type of studies available, a scoping review methodology was chosen (18). The study protocol (published on OFS, DOI: 10.17605/OSF.IO/STQ9D) complies with the ‘Preferred Reporting Items for Systematic Reviews and Meta-Analyses for Scoping Reviews (PRISMA-ScR) checklist’ (19). The following inclusion criteria were applied: Papers needed to respect the categories of the PICo-framework (**P**roblem: malaria situation; **I**nterest: the public health impact of the COVID-19 pandemic; **Co**ntext: sub-Saharan Africa) (20). No restrictions regarding the study type and the publication status were applied. Possible languages were English, French and German; papers published between December 1, 2019, and June 9, 2021, were included. In line with the protocol, the search strategy was developed, and the following databases were searched: PubMed; Ovid MEDLINE(R); Web of Science; Biosis Previews; MedRxiv, and The Lancet.

Grey literature was included using WHO database and Google Scholar. Three broad blocks of search terms were used: (1) COVID-19, (2) malaria, (3) sub-Saharan Africa. The detailed search strategy is available in appendix 1.

For the extracted findings two researcher (OM and AH) conducted independently the title screening, then the abstract screening and finally the full text review. The papers selected for full-text reading were assessed for eligibility; ineligible papers did not include information on public health relevant consequences of the COVID-19 pandemic on malaria in SSA. Inclusion decisions depended on whether the paper agreed to the PICo-framework and the formal eligibility criteria. Results were compared after each step for discussion and for reaching a consensus. For the analysis of the finally included papers, a data extraction table was constructed (appendix 2).

The following information was extracted from the papers: Authors, title, study place, study population, study design and outcome. Moreover, the papers were categorized by study type: modeling study, report (country report, general report, case report), review, opinion paper, and policy guideline. The information content was structured and analyzed around the following themes:

- Modeled impact of COVID-19 on malaria
- Diagnostic and clinical aspects
- Access to health care services
- Availability of curative and preventive malaria commodities
- Impact on malaria programs
- Epidemiologic data from countries

Based on these findings, a conceptual framework was created, with input from all co-authors (figure 1).

**Figure 1:**
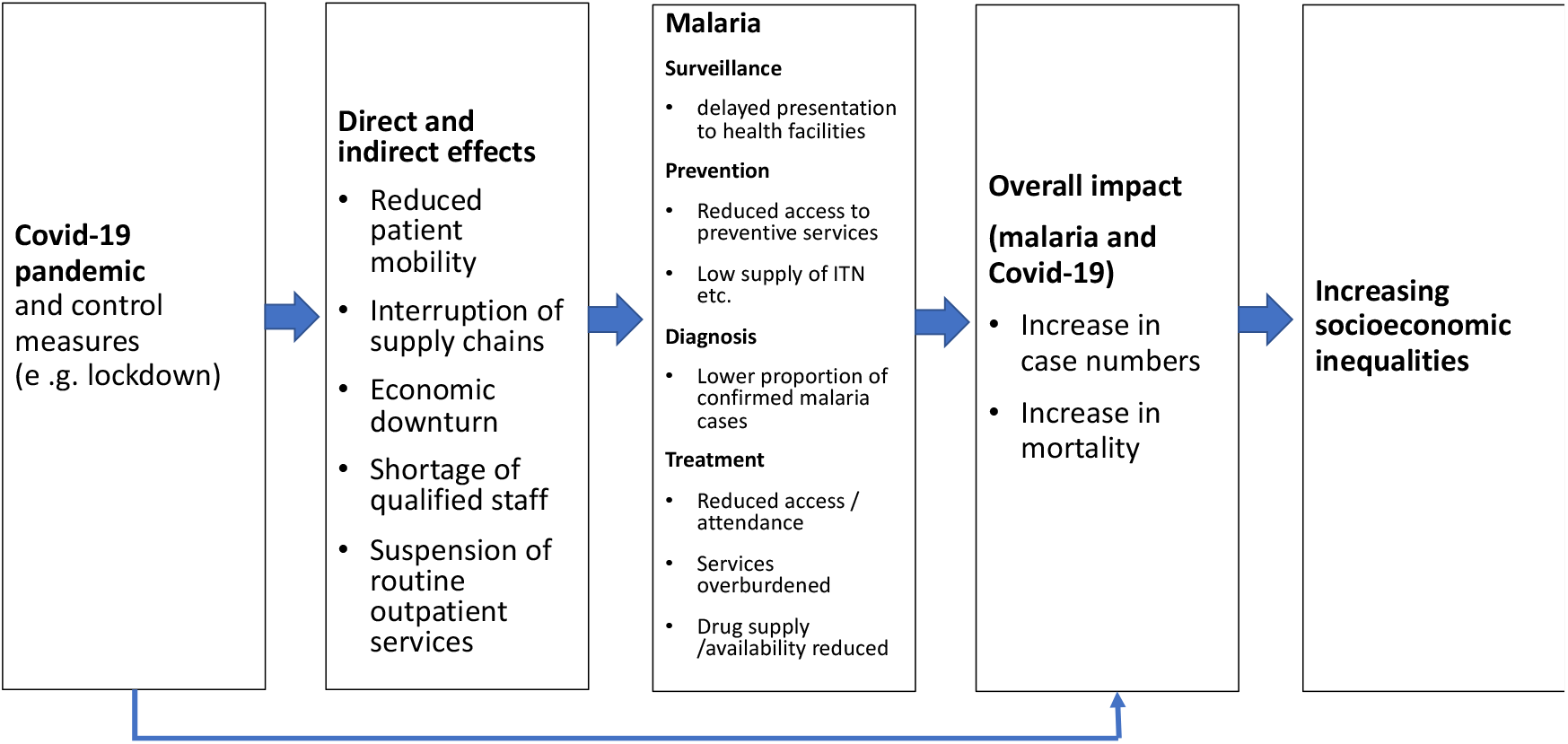
Conceptual framework presenting the different factors resulting from the global COVID-19 pandemic on the malaria situation in SSA.

## Results

Figure 2 visualizes the study selection process. The initial search produced 851 documents. After removal of 203 duplicates, 648 documents underwent title and abstract screening. After exclusion of 535 documents which did not meet the inclusion criteria, 113 papers were included for full text review. 9 papers were added from reference screening; 71 were excluded as they also did not meet the inclusion criteria. Thus, a total of 51 papers were reviewed (6 modeling studies, 10 country reports, 6 general reports, 1 case report, 9 review papers, 18 opinion papers, and 1 policy guideline).

**Figure 2:**
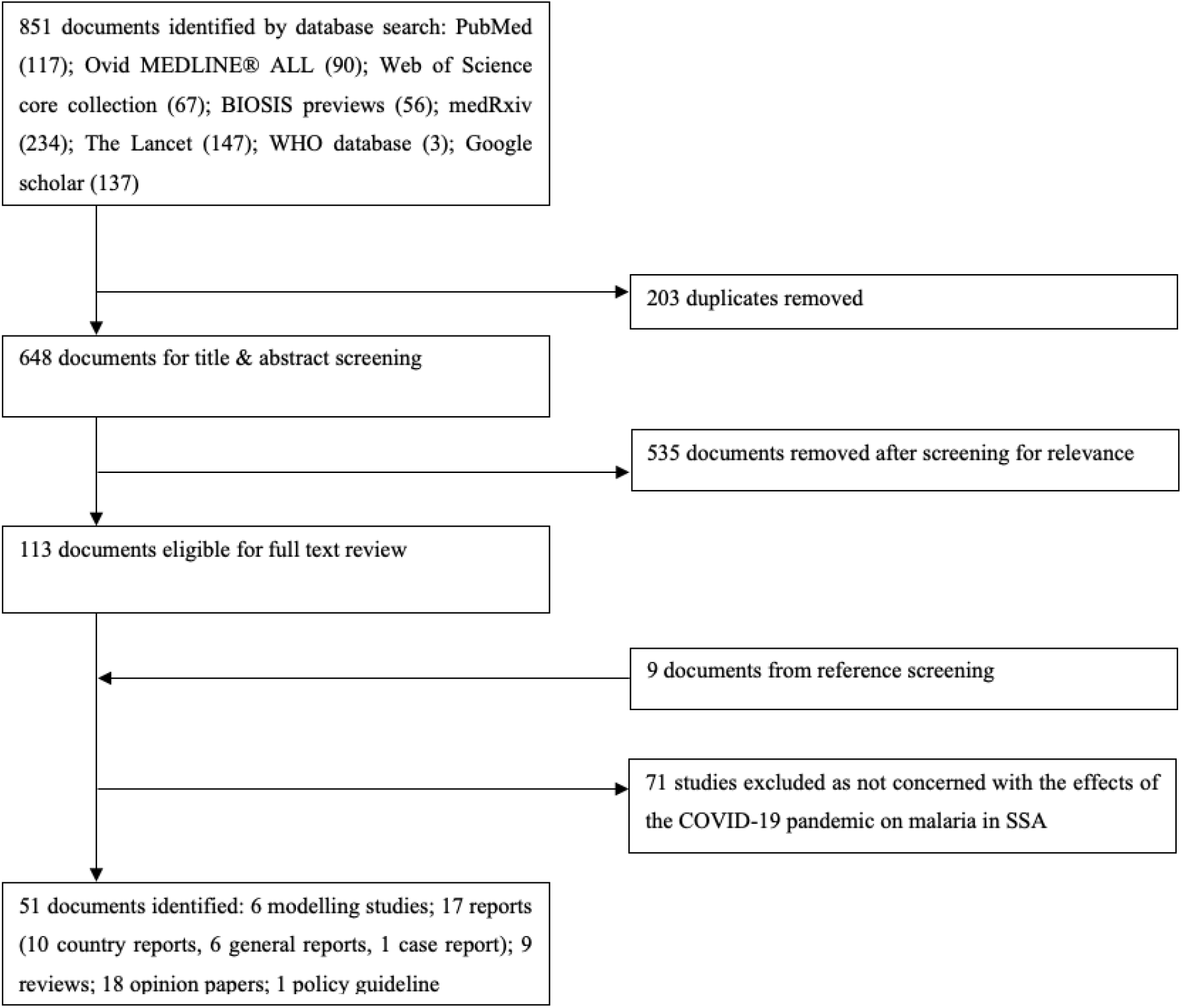
Study selection process

### Modeled impact of COVID-19 on malaria

Five papers predicted the evolution of the malaria burden in SSA based on different potential scenarios. Considering primarily a reduced access to effective antimalarial treatment and reduced insecticide-treated mosquito net (ITN) distribution, Weiss et al. predicted in their worst-case scenario (75% less anti-malarial drugs and ITNs) and for the year 2020 that in SSA countries malaria cases would increase by 22% (from 215 to 262 million) and malaria deaths by 99% (from 386,000 to 769,000); the lower access to antimalarial treatment had a larger effect than reduced ITN distribution (21). These estimates mirror those by the WHO, but the authors described the effects of nine different scenarios compared to the effects of three scenarios by Weiss et al. (22). Comparable estimates were published by Sherrard-Smith et al.; for the scenario of complete interruption of ITN distribution and 50% decreased access to antimalarials; they predicted malaria deaths would increase in SSA to 779,000 for the year 2020 (23). A further analysis by WHO predicted up to 100,000 additional deaths in 2020 with a 50% lower access to antimalarials (17). However, all these authors emphasized that the projected effects on malaria services and mortality are highly uncertain because these estimates are heavily dependent on how countries respond to the COVID-19 pandemic. Regarding the relative burden of COVID-19 in Africa, one study concluded that the excess Disability-Adjusted Life Years (DALYs) lost by malaria due to COVID-19 may exceed those directly lost due to COVID-19 (24).

### Diagnostic and clinical aspects

The clinical manifestations of COVID-19 and malaria largely overlap; fever, headache, joint pain, respiratory symptoms, and general weakness are frequently seen with both diseases (25-27). Thus, diagnosis based on symptoms alone can result in inadequate treatment, with potentially harmful consequences. Untreated malaria can be rapidly fatal and COVID-19 patients must be quarantined to interrupt community transmission (14, 28). Despite increasing availability of rapid diagnostic tests (RDTs) for malaria in all endemic areas, presumptive diagnosis of malaria is still common in SSA, and the WHO Malaria Technical Guidelines adapted to COVID-19 confirm this situation (29, 30). Initial information available for 2020 suggests major disruptions in malaria diagnosis and treatment due to COVID-19 (31, 32).

Human travel history is important for SARS-CoV-2 and malaria, as for both of them asymptomatic persons can spread and/or maintain transmission of the infectious agent (26). Malaria might have been reduced by the COVID-19 movement restrictions, especially in heterogenous malaria-endemic settings where transmission frequently results from migration flows of infected individuals across different regions (33). Moreover, malaria and SARS-CoV-2 co-infections may be associated with clinical disease modification, although data on this are limited (27, 34-36).

While symptomatic malaria affects mainly children and younger age groups in endemic areas, COVID-19 affects all age groups but is more frequently symptomatic and severe with increasing age (34). However, in areas of low malaria transmission, the age groups affected by the two diseases largely overlap (37). RDTs are essential for malaria diagnosis in rural SSA and may also become important for COVID-19, as the PCR test capacity is very limited (32). However, the impact of rather low sensitivity and specificity of COVID-19 RDTs is still under intense discussion (27, 38). An additional challenge for differential diagnosis is the increasing frequency of gene-mutated *Plasmodium* parasites, especially in the Horn of Africa, that escape detection by standard RDTs (37).

The role of antimalarials, e.g. artemisinin derivates and chloroquine (CQ), in the COVID-19 pandemic is complex. Various artemisinin derivates, artemisinin-based combination therapies (ACTs) as well as CQ have been shown to be effective against SARS-CoV-1 and SARS-CoV-2 *in vitro* (39-42). However, such beneficial effect was not confirmed by several clinical trials (43-46). The wide use of these treatments in highly malaria endemic countries has been suggested to be responsible for the reported low COVID-19 burden in SSA (36, 47). On the other hand, the increased usage of these drugs for COVID-19 prevention and treatment in some malaria endemic countries might have reduced malaria (25). A frequent off-label use of artemisinin-based drugs may also increase the likelihood of emerging drug resistance and thus threatens the most important of the remaining effective antimalarials (33, 48-50).

### Access to health care services

The COVID-19 pandemic in SSA endangers access to health care services due to several factors. Direct factors include restricted services and closures of health facilities because of reduced health care worker (HCW) capacity due to lack of personal protective equipment (PPE), stigmatization, fear of getting infected, or absence due to COVID-19 quarantine, disease or death (27, 32, 50, 51). Delayed treatment results in prolonged gametocyte carriage and additional opportunities for transmission.

Moreover, because of overload of COVID-19 patients and consequently reduced time to manage other diseases, or due to movement and travel restrictions and for fear of becoming infected with COVID-19, sick individuals with diseases other than COVID-19 do no longer attend health facilities (33, 48, 52). As older people fear severe COVID-19 disease and may thus avoid visiting health facilities, this might affect children the most as they depend on their care givers if sick, including for malaria (35, 49). Stay-at-home advices for febrile diseases, especially at the beginning of the pandemic, enhanced such a behavior (17, 33).

Indirect factors include reduced income during lockdowns due to inability to perform informal work, and subsequently reduced purchasing power (52). The resulting increase in poverty leads to challenges for paying the costs for routine care, drugs, or transportation fees (51). Lockdowns, movement restrictions and border closures further complicate access to health facilities and have also threatened the functioning of malaria surveillance systems (16, 28, 33, 51). Institutional mistrust and lack of valid information further reduced visits to health care facilities and reduced uptake of preventive measures; as an example, myths about the spread of COVID-19 via ITNs led to a reduced usage of this essential intervention in Sierra Leone (25).

### Availability of curative and preventive commodities and medicines

Increased material costs, reluctance of producers to invest, travel restrictions, border closures, and lockdowns resulted in a lower availability of medical malaria products (26, 28, 48). Low- and middle-income countries (LMICs) are disproportionately affected as they essentially rely on importation of these commodities (52). Excessive use of antimalarials for COVID-19 prevention and treatment in some regions has led to shortages for their original purpose (17, 30). Some international companies switched from the production of malaria products to COVID-19 products (48, 49, 51). Difficult access to health facilities lowered the availability of essential drugs and increased their price, with subsequent increases in purchase and usage of sub-standard drugs and alternative medicines (28, 51-53). In addition, PPE needed for the implementation of different malaria services (e.g. indoor residual spraying of insecticides, IRS) has become scarce and expensive on global markets (17, 54).

### Impact of the pandemic on malaria programs

The extent of the pandemic’s impacts on malaria depends on the timing of its waves. The largest effects may occur if the COVID-19 transmission peaks and the planned malaria campaigns overlap (21, 23, 53, 55). About three quarters of malaria-affected countries reported disruptions of malaria services and programs (17, 32, 33, 50, 53, 56-58). Reallocation of funds from other disease control programs to the control of COVID-19 have been common and pose great problems for malaria control (30, 32, 35, 59, 60). Ongoing malaria programs (e.g. IRS, ITN interventions) need to be adapted to the restrictions associated with COVID-19 control measures, which requires additional financial resources (32, 33, 36). Programs for vulnerable populations living in remote areas are particularly at risk as they strongly depend on logistics and external financing (33, 48). Disrupted ITN programs will lead to increased malaria transmission as 80% of the nets are distributed through mass campaigns (22, 48, 53, 55). IRS campaigns face many challenges as they require direct household contact (33, 50, 57).

Nevertheless, these challenges have led to new approaches: Benin digitalized its ITN mass distribution campaign using a ‘no touch’ payment for campaign workers. The national strategy was changed from a fixed-point to a door-to-door-distribution procedure, which enabled health workers to provide additional community health education on COVID-19 and other aspects; other countries followed the Benin model and by the end of 2020, 90% of all globally planned malaria prevention campaigns had been implemented (17, 28, 54, 57, 59).

### Epidemiologic data from countries

Compared to previous years, fewer papers provided data from African countries on the actual number of malaria cases and deaths in 2020. A small study from Sierra Leone reported a significant lower number of malaria outpatient visits in one health facility during the March/April 2020 lockdown period as compared to the same period in 2019 (29). In addition, preliminary national data from Uganda point to a reduction of malaria cases diagnosed in health facilities during the first quarter of 2020 compared to the same period in 2019 (61). Another study from Uganda reported a 54% decrease in visits for malaria treatment of febrile children; visits for antenatal care declined by 26%, restricting the delivery of intermittent preventive malaria treatment in pregnancy (IPTp) (62). In the Democratic Republic of the Congo (DRC), lower attendance to health facilities for malaria treatment ranged from 20% to 90%, depending on local lockdown measures (63). In contrast, a study from one rural district in Zimbabwe reported a large increase in malaria cases in 2020 compared to previous years, which was associated with delayed IRS in 2020 (50). These findings were confirmed by national data from Zimbabwe, which compared the number of malaria cases and deaths in 2020 with those in previous years; in 2020, there was a large excess of reported malaria cases and deaths (27, 64). Moreover, national data from Zambia showed an increase of malaria cases between August 2019, and June 2020; however, no data from control periods were provided (65).

## Discussion

The COVID-19 pandemic has a massive impact on nearly all countries across the world. While the initial spread of SARS-CoV-2 to Africa has been slow and the COVID-19 burden appears to be much lower than in other continents, the pandemic carries a high potential to negatively affect the control of other diseases such as malaria (7). It has already been shown, that the pandemic has resulted in major reductions of the incidence of other respiratory diseases due to various effects (66). Moreover, it has been predicted that the pandemic will result in major disruptions of routine childhood vaccinations, which may cause an increase in vaccine-preventable infectious diseases in SSA (67). Both malaria and COVID-19 affect disproportionally the low socio-economic classes (28, 32, 68). It is possible that the COVID-19 pandemic and its indirect effects, including the measures to contain it, may produce collateral damages similar to those seen six years ago during the West African Ebola epidemic, i.e. a sharp increase of malaria deaths which finally exceeded the direct Ebola mortality (17, 36, 69). Thus, understanding how the COVID-19 pandemic affects malaria control measures is of extreme importance for SSA (17, 59).

Accelerated malaria control efforts since the early 21^st^ century have significantly reduced the malaria burden in Africa and worldwide (17). Control strategies include ITN and IRS interventions, early diagnosis and rapid treatment with ACT, and intermittent preventive treatment for infants, children and pregnant women (14). However, the rate of reduction in malaria morbidity and mortality in SSA has recently stalled, and the initial overall positive trend could be seriously reversed due to the effects of the COVID-19 pandemic as shown in several modelling studies (17, 21-23).

In accordance with our conceptual framework, four major themes likely play a major role for the effects of the COVID-19 pandemic on malaria in SSA: (1) diagnostic and clinical aspects; (2) access to health care services; (3) availability of curative and preventive malaria commodities; and (4) impact on malaria prevention programs. While diagnostic and clinical aspects will play an obvious role due to the overlapping symptoms of both diseases (27, 70, 71), therapeutic aspects related to initial misperceptions regarding the efficacy of certain antimalarials against COVID-19 may have been overemphasized (25, 36). Co-infection with malaria may complicate COVID-19, while immunomodulation caused by previous malaria exposure may result in less severe COVID-19, as previously also shown in other respiratory diseases (72-75). Reduced access to health care services due to direct and indirect effects of the pandemic has a negative impact on access to antimalarial treatment, thus it would likely have a major effect on the malaria burden in endemic countries (17, 49, 53). This will be compounded by the clear negative impact of the pandemic on global supply chains for curative and preventive malaria commodities (48, 52). The consequences of the pandemic for preventive malaria control programs have been much emphasized by many of the reviewed papers and particularly in modeling papers. However, as an effect of such early warnings, country programs and funding for malaria have probably adapted rapidly to the pandemic as early as 2020, which may have reduced the modeled impact (28, 59). International actors like the WHO may have contributed to the prevention of some worst-case scenarios by providing adapted malaria strategies and keeping malaria in their priorities (17, 30).

Until June 2021, only a few reports provided actual epidemiological data on malaria in SSA during the first wave of the pandemic in 2020, thus drawing conclusions on these data might be premature. However, these reports showed that the number of reported malaria cases in Sierra Leone, Uganda and the DRC, what are more highly malaria endemic countries, was much lower than expected (29, 61-63), while the number of reported malaria cases in Zimbabwe and Zambia, that are countries of low endemicity, was higher than in previous years (27, 29, 64, 65). It could be speculated that possibly lower access to health care services in combination with impaired malaria surveillance systems may have led to a lower number of reported malaria cases and deaths in these selected highly endemic countries. In the two low endemic southern SSA countries, disruption of malaria control activities within relatively well-functioning health systems, including surveillance activities, may have resulted in a higher number of reported malaria cases and deaths. More information from other African endemic countries is needed to fully assess such developments (59, 76). As the COVID-19 pandemic is far from being under control in most LMICs as new and more infectious SARS-CoV-2 variants are emerging, and as SSA countries have limited access to COVID-19 vaccines, dramatic increases of the malaria burden may occur (59, 77, 78). Although the findings of existing modeling studies are already alarming, the final impact of the pandemic on the malaria burden could be even more devasting (21, 51). Better education, sensitization and de-stigmatization of both diseases is essential, including emphasis on early care seeking behaviour, which needs also more community participation (25, 29). Community health workers should be encouraged to treat all uncomplicated malaria cases in the community and to refer to health facilities only severe cases (51, 79). As 2020 was a year with many planned malaria prevention campaigns, the negative effects of disrupted programs would probably last for some years (21, 23). Fortunately, the international community, including the WHO, acted fast to counteract such developments (17). However, there is the need for more support for SSA countries from the international community and from high income countries (32). Malaria, one of Africa’s deadliest diseases which disproportionally affects the most vulnerable population groups, must be kept under control (16, 35, 59).

## Conclusion

The findings from this review provide evidence for a significant but diverse impact of the COVID-19 pandemic on the malaria burden in SSA. Only results of further studies will enable a full understanding of these developments and its public health consequences. In the meantime, SSA countries need more support from the international community including the urgent delivery of COVID-19 vaccines for high-risk groups.

## Data Availability

The datasets used and/or analysed during the current study are available from the corresponding author on reasonable request.

## List of abbreviations

ACT: artemisinin-based combination therapy
COVID-19: coronavirus disease 2019
CQ: chloroquine
DRC: Democratic Republic of the Congo
HCW: health care worker
IPTp: intermittent preventive treatment in pregnancy
IRS: indoor residual spraying of insecticides
ITN: insecticide-treated mosquito net
LMIC: low- and middle-income country
PICo: problem, interest, context
PPE: personal protective equipment
SARS-CoV-2: severe acute respiratory coronavirus type 2
SSA: sub-Saharan Africa
WHO: World Health Organization

## Declarations

### Ethics approval and consent to participate

Not applicable

### Consent for publication

Not applicable

### Authors’ contributions

AH and OM performed the systematic search and screening. AH wrote the first draft, GL did the methodological foundation, OR drafted the conceptual framework; all authors read, reviewed and approved the final manuscript.

## Acknowledgements

Not applicable

## Appendix

## Appendix 1: Detailed search strategy

*Concept 1:* COVID-19

“COVID-19”[ALL] OR “COVID*”[ALL] OR “SARS-CoV-2”[ALL] OR “coronavirus*”[ALL] OR “2019-nCoV disease”[ALL] OR “betacoronavirus”[ALL] OR “nCoV”[ALL] OR “COVID-19” [Supplementary Concept] OR “severe acute respiratory syndrome coronavirus 2”[nm]

*Concept 2:* malaria

“malaria*”[ALL] OR “paludism*”[ALL] OR “Malaria”[Mesh] OR “Malaria/prevention and control”[MAJR]

*Concept 3:* sub-Saharan Africa

“africa”[ALL] OR “sub-saharan”[ALL] OR “SSA”[ALL] OR “south of the sahara”[ALL] OR “Africa South of the Sahara”[Mesh]

*Concept 1 AND concept 2 AND Concept 3*

| Search number | Query | Sort By | Filters | Search Details | Results | Time |
| --- | --- | --- | --- | --- | --- | --- |
| 5 | #1 AND #2 AND #3 | Most Recent | from 2019/12/1 - 2021/6/9 | ((("COVID-19"[All Fields] OR "covid*" [All Fields] OR "SARS-CoV-2"[All Fields] OR "coronavirus*" [All Fields] OR "2019-nCoV disease"[All Fields] OR "betacoronavirus"[All Fields] OR "nCoV"[All Fields] OR "COVID-19"[Supplementary Concept] OR "severe acute respiratory syndrome coronavirus 2"[Supplementary Concept]) AND ("malaria*" [All Fields] OR "paludism*" [All Fields] OR "Malaria"[MeSH Terms] OR "malaria/prevention and control"[MeSH Major Topic]) AND ("africa"[All Fields] OR "sub-saharan"[All Fields] OR "SSA"[All Fields] OR "south of the sahara"[All Fields] OR "Africa South of the Sahara"[MeSH Terms])) AND (2019/12/1:2021/6/9[pdat]) | 117 | 13:34:11 |
| 4 | #1 AND #2 AND #3 | Most Recent |  | ("COVID-19"[All Fields] OR "covid*" [All Fields] OR "SARS-CoV-2"[All Fields] OR "coronavirus*" [All Fields] OR "2019-nCoV disease"[All Fields] OR "betacoronavirus"[All Fields] OR "nCoV"[All Fields] OR "COVID-19"[Supplementary Concept] OR "severe acute respiratory syndrome coronavirus 2"[Supplementary Concept]) AND ("malaria*" [All Fields] OR "paludism*" [All Fields] OR "Malaria"[MeSH Terms] OR "malaria/prevention and control"[MeSH Major Topic]) AND ("africa"[All Fields] OR "sub-saharan"[All Fields] OR "SSA"[All Fields] OR "south of the sahara"[All Fields] OR "Africa South of the Sahara"[MeSH Terms]) | 121 | 13:33:52 |
| 3 | "africa"[ALL] OR "sub-saharan"[ALL] OR "SSA"[ALL] OR "south of the sahara"[ALL] OR "Africa South of the Sahara"[Mesh] | Most Recent |  | "africa"[All Fields] OR "sub-saharan"[All Fields] OR "SSA"[All Fields] OR "south of the sahara"[All | 372,746 | 13:33:40 |
|  |  |  |  | Fields] OR "Africa South of the Sahara"[MeSH Terms] |  |  |
| 2 | "malaria*"[ALL] OR<br>"paludism*"[ALL] OR<br>"Malaria"[Mesh] OR<br>"Malaria/prevention and control"[MAJR] | Most Recent |  | "malaria*"[All Fields] OR<br>"paludism*"[All Fields] OR<br>"Malaria"[MeSH Terms] OR<br>"malaria/prevention and control"[MeSH Major Topic] | 104,127 | 13:33:32 |
| 1 | "COVID-19"[ALL] OR<br>"COVID*"[ALL] OR "SARS-CoV-2"[ALL] OR "coronavirus*"[ALL] OR<br>"2019-nCoV disease"[ALL] OR<br>"betacoronavirus"[ALL] OR<br>"nCoV"[ALL] OR "COVID-19"[Supplementary Concept] OR "severe acute respiratory syndrome coronavirus 2"[nm] | Most Recent |  | "COVID-19"[All Fields] OR<br>"covid*"[All Fields] OR "SARS-CoV-2"[All Fields] OR<br>"coronavirus*"[All Fields] OR<br>"2019-nCoV disease"[All Fields] OR<br>"betacoronavirus"[All Fields] OR<br>"nCov"[All Fields] OR "COVID-19"[Supplementary Concept] OR<br>"severe acute respiratory syndrome coronavirus 2"[Supplementary Concept] | 158,835 | 13:33:26 |

## Appendix 2 Data extraction table

*Table of included studies*

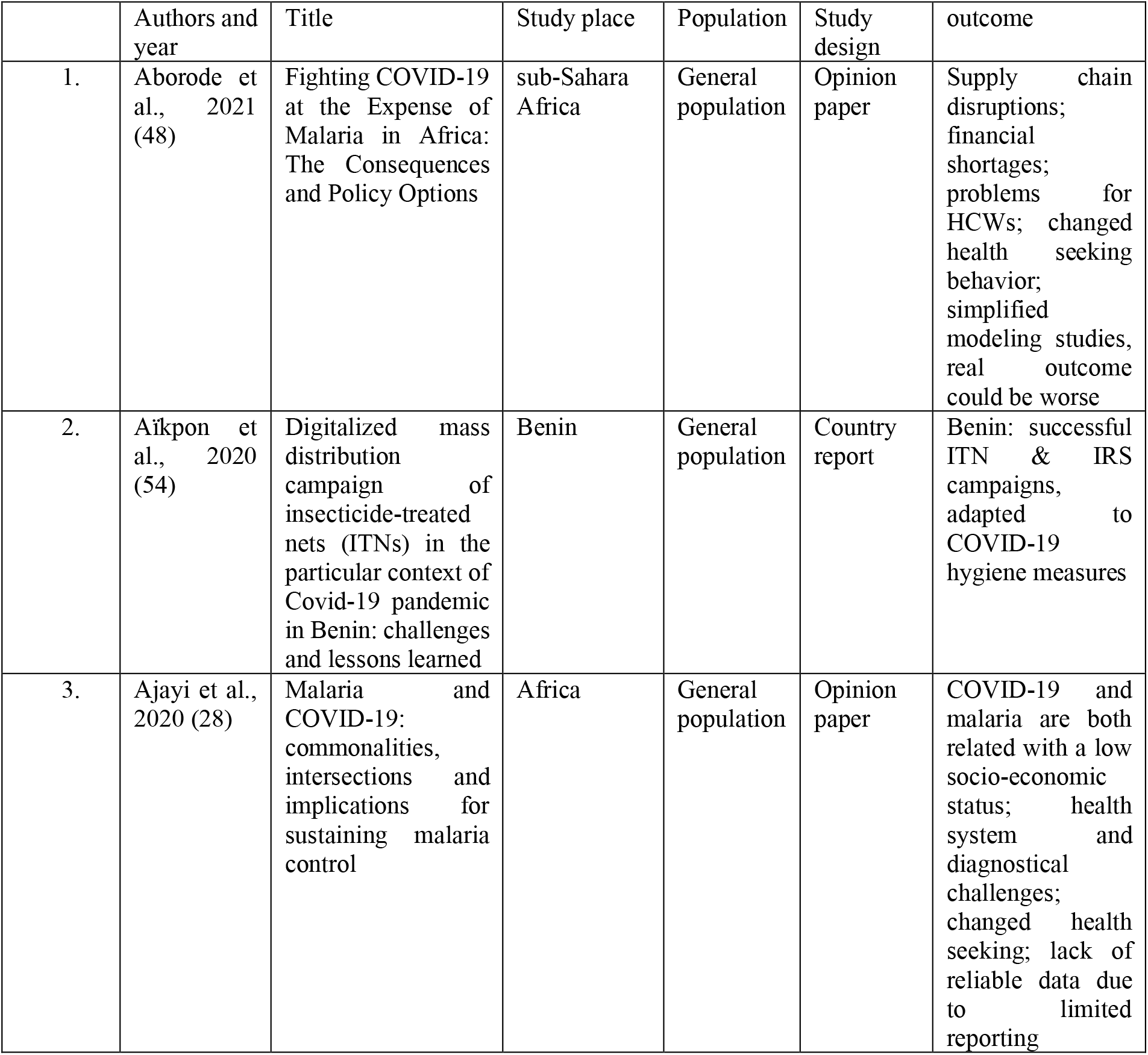
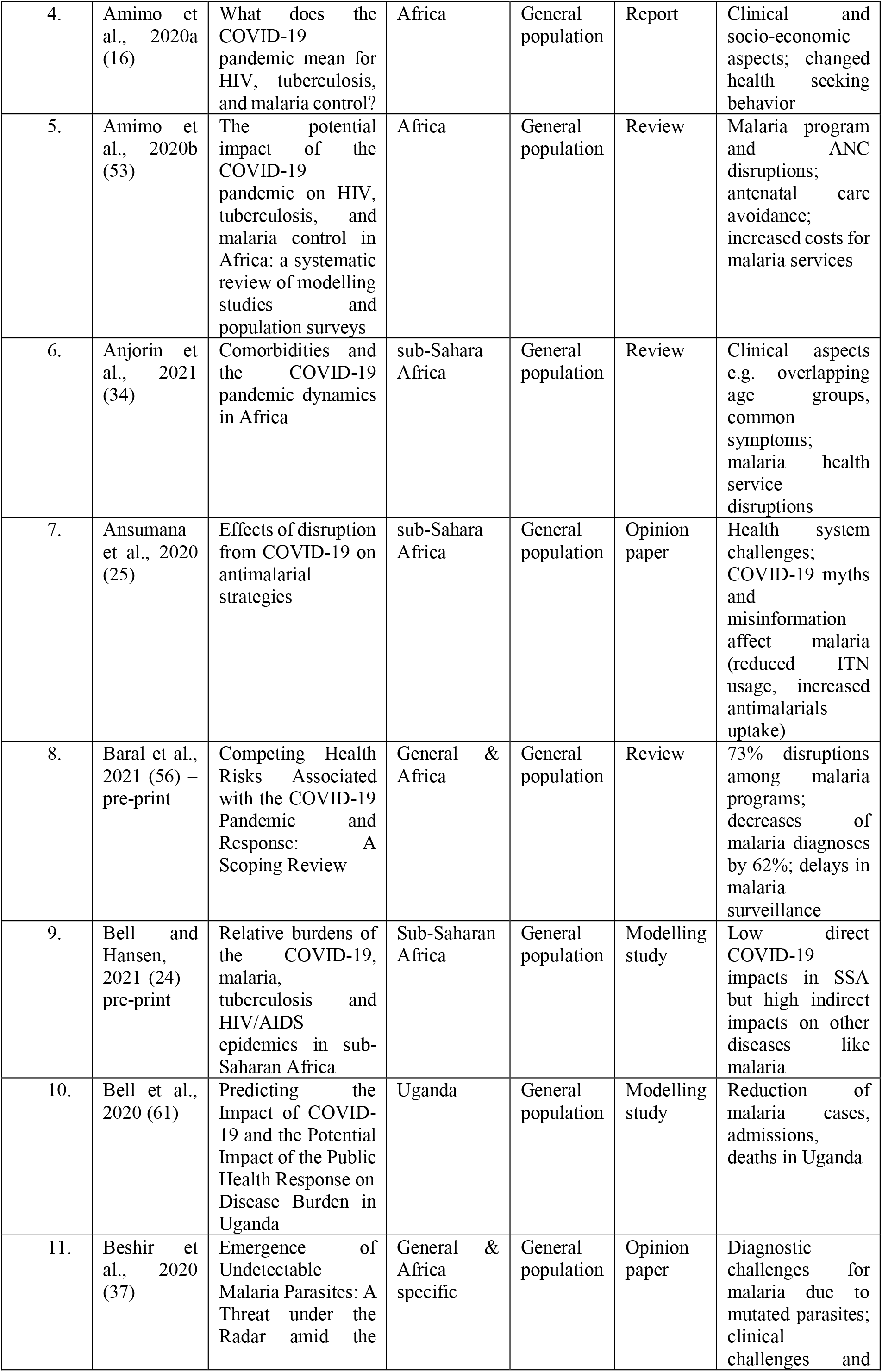
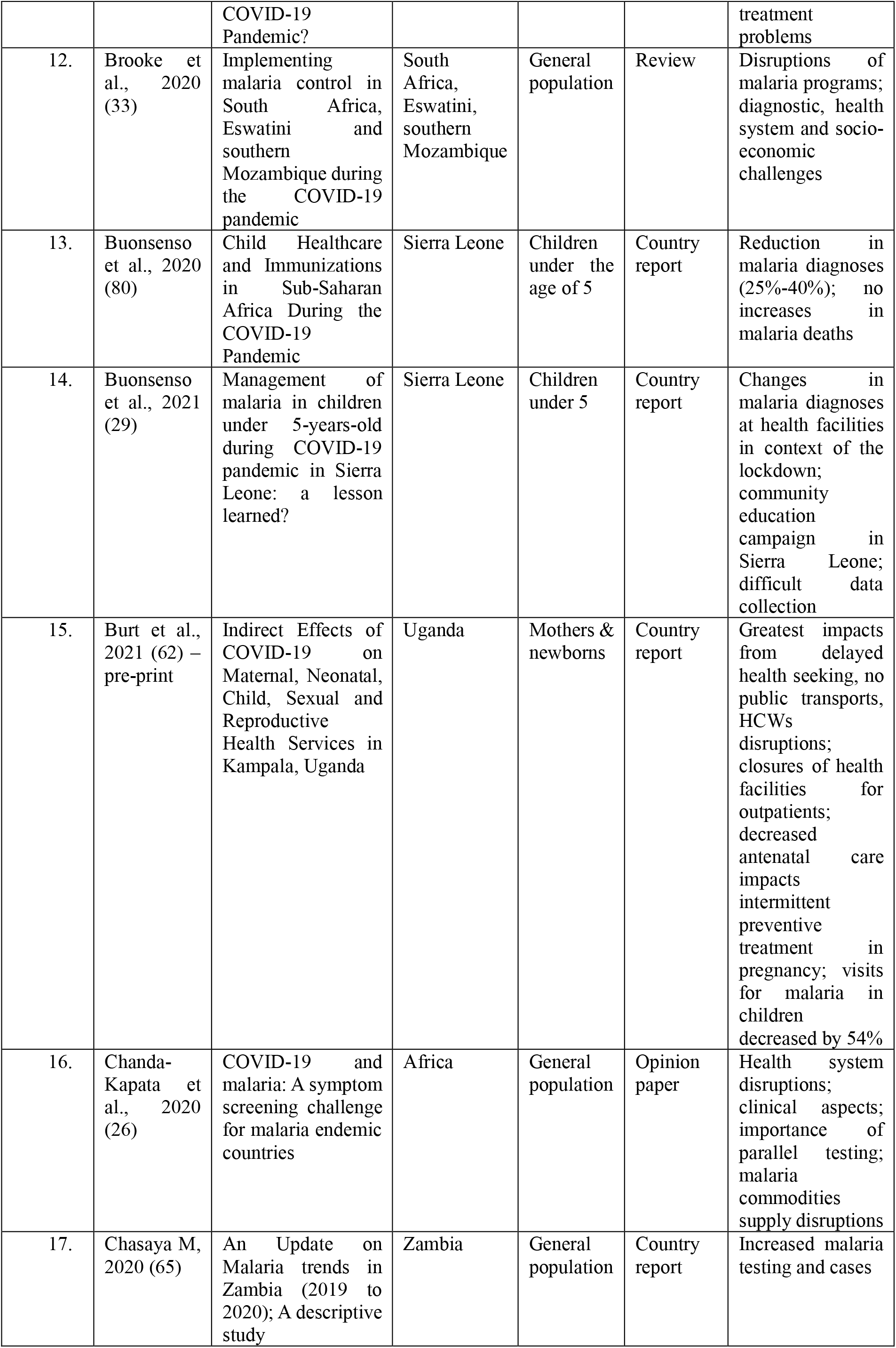
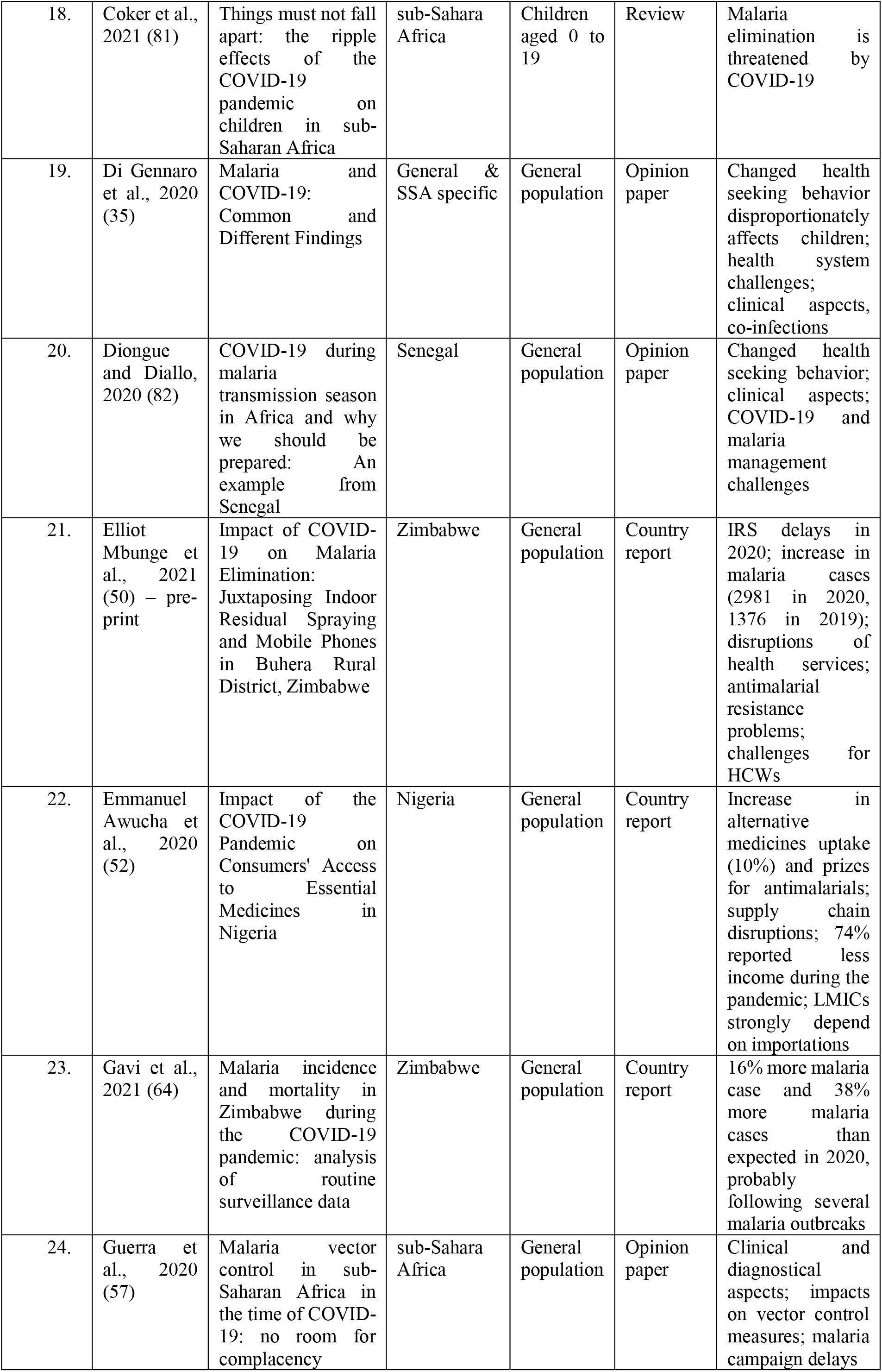
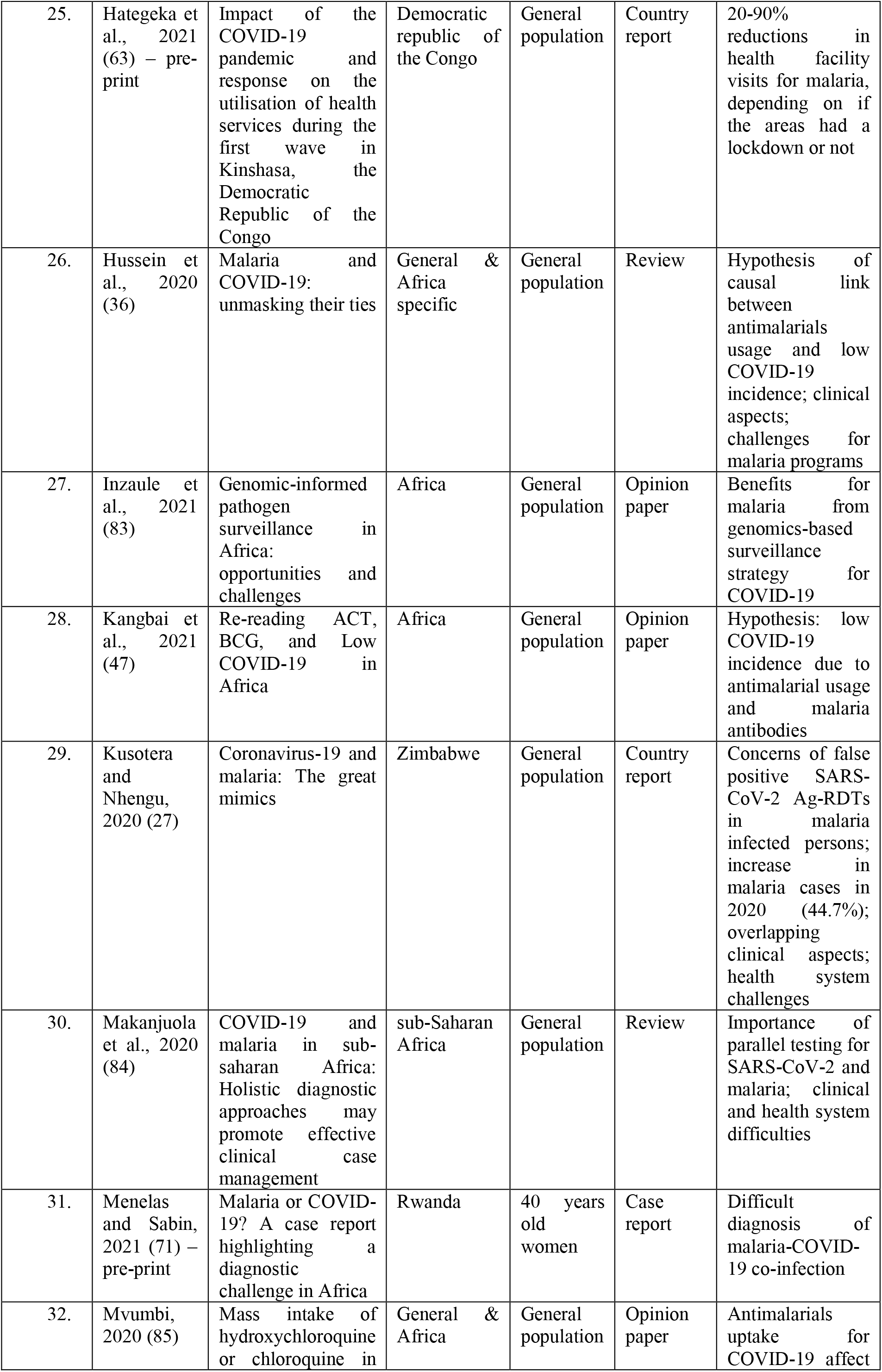
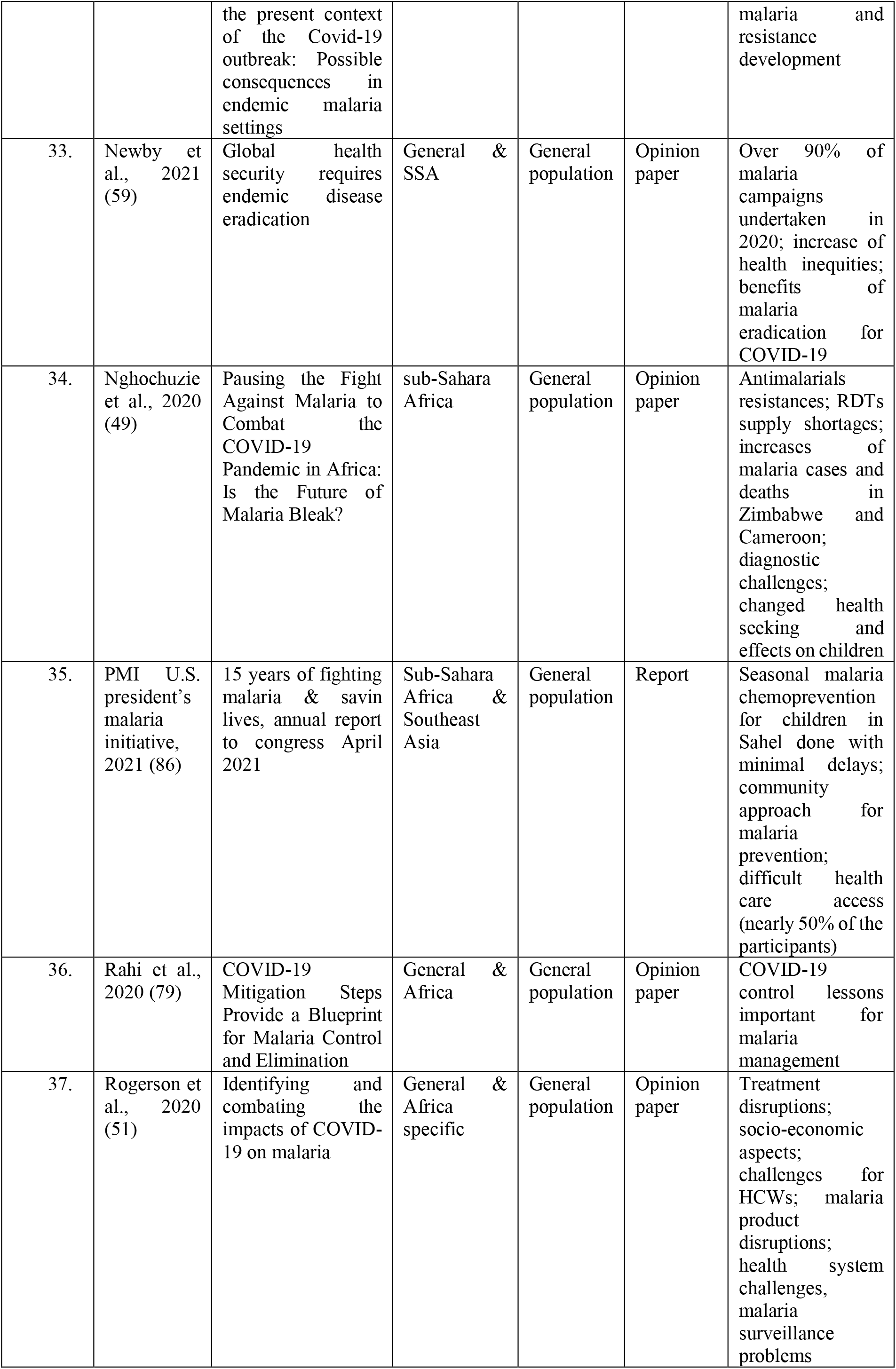
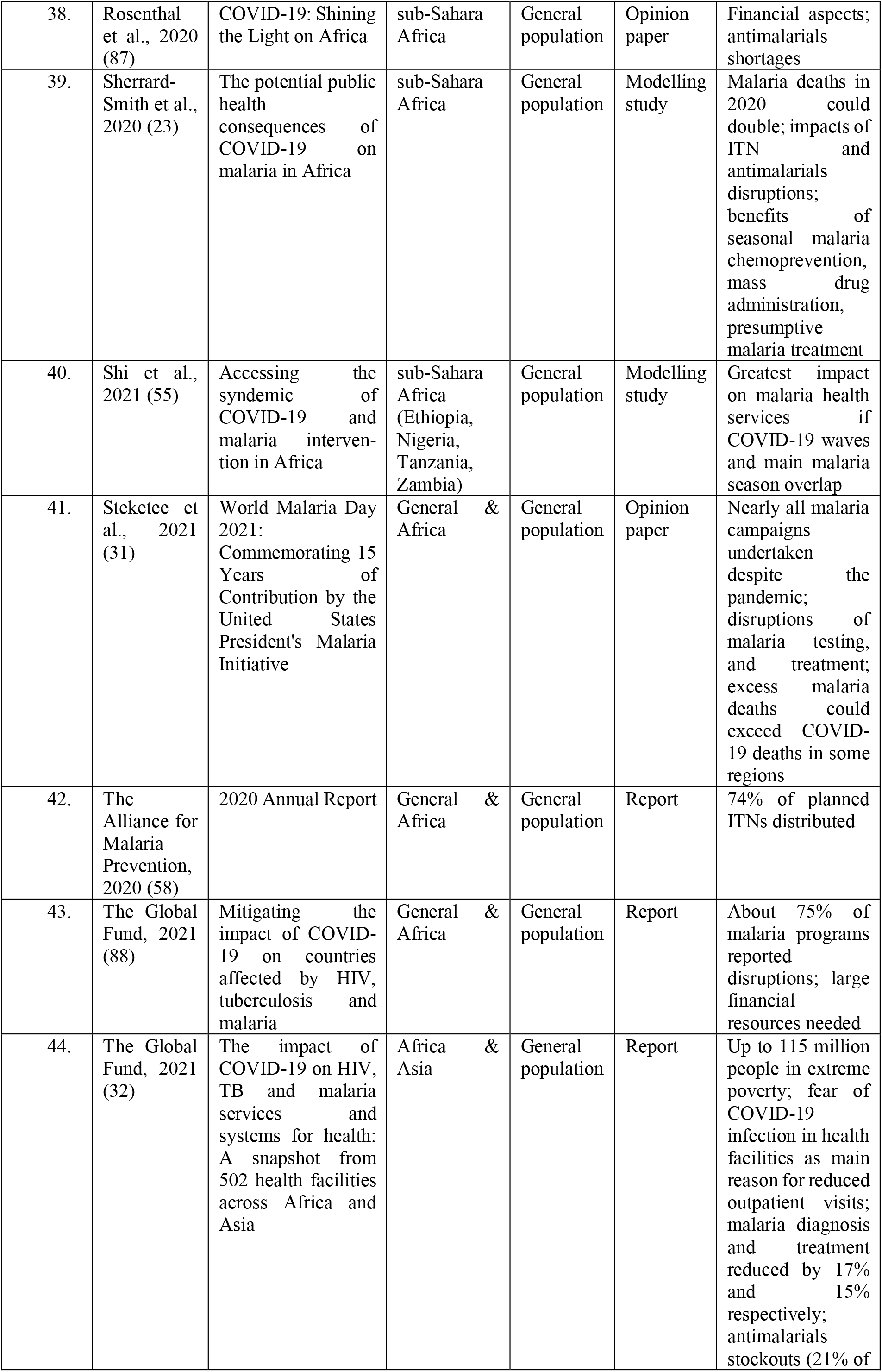
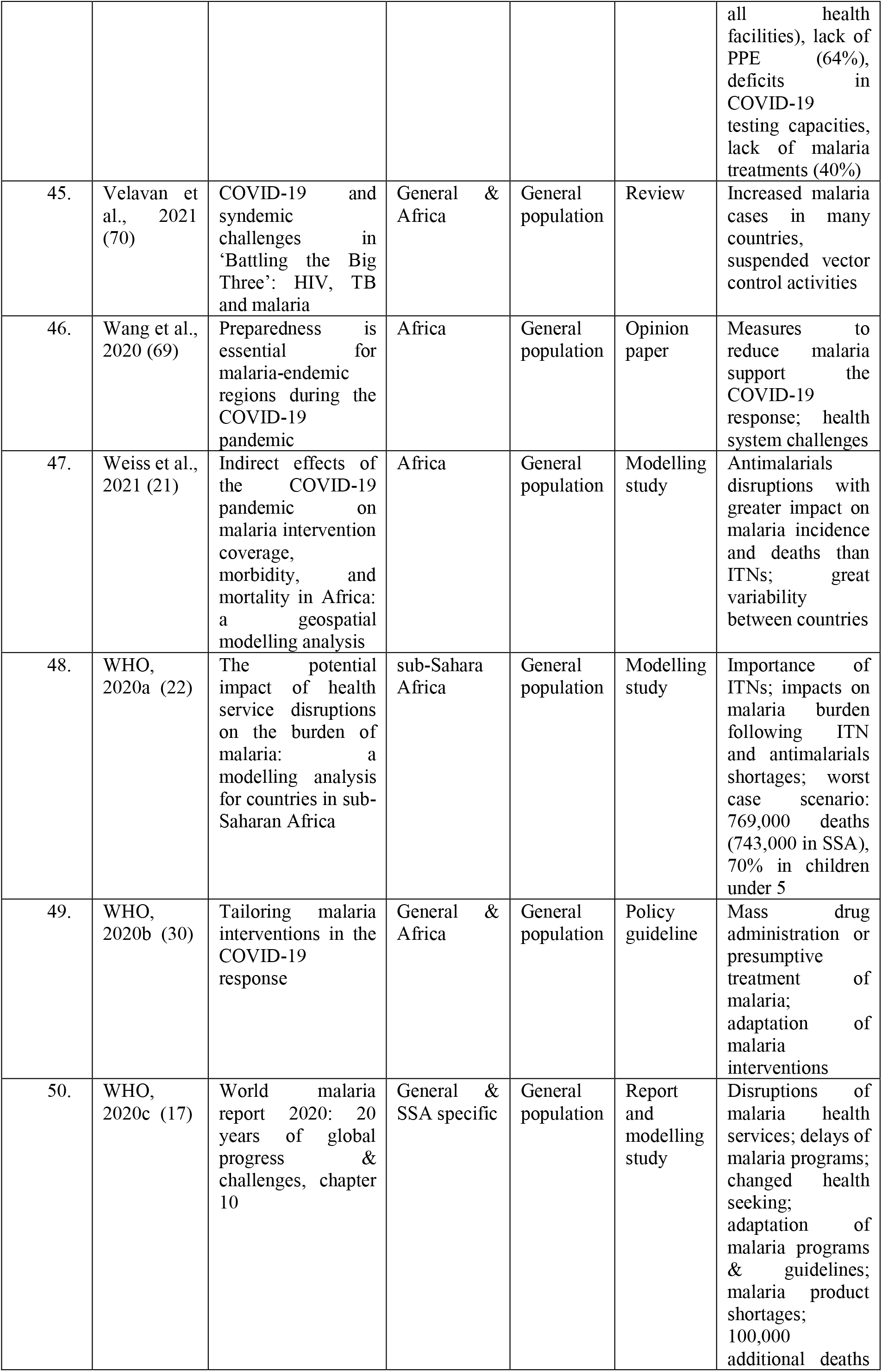
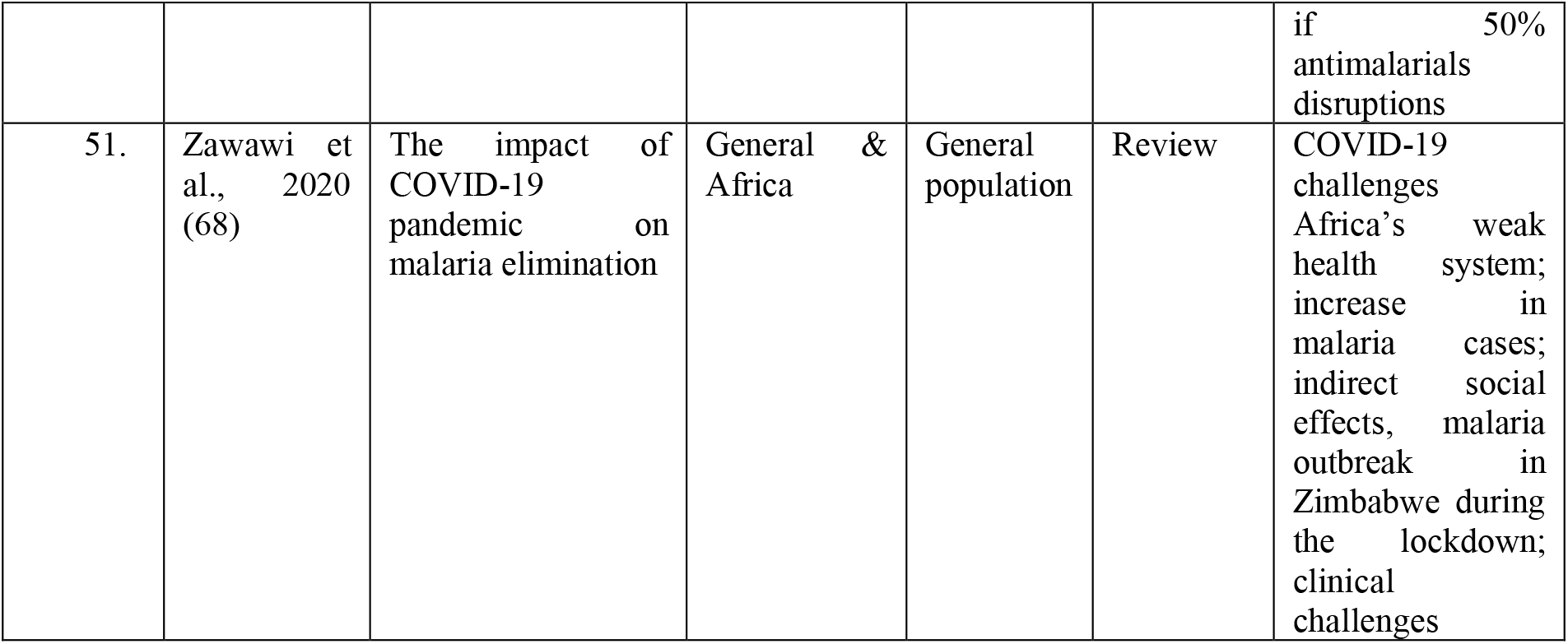

## Appendix 3

## Bibliography

1. Dawood FS, Ricks P, Njie GJ, Daugherty M, Davis W, Fuller JA, et al. Observations of the global epidemiology of COVID-19 from the prepandemic period using web-based surveillance: a cross-sectional analysis. The Lancet Infectious Diseases. 2020;20(11):1255–62.

2. Müller O, Lu G, Jahn A, Razum O. COVID-19 Control: Can Germany Learn From China? Int J Health Policy Manag. 2020;9(10):432–5.

3. WHO Coronavirus (COVID-19) Dashboard [Internet]. World Health Organization,. 2021. Available from: https://covid19.who.int/table.

4. Lu G, Razum O, Jahn A, Zhang Y, Sutton B, Sridhar D, et al. COVID-19 in Germany and China: mitigation versus elimination strategy. Glob Health Action. 2021;14(1):1875601.

5. Maeda JM, Nkengasong JN. The puzzle of the COVID-19 pandemic in Africa. Science. 2021;371(6524):27–8.

6. Massinga Loembé M, Tshangela A, Salyer SJ, Varma JK, Ouma AEO, Nkengasong JN. COVID-19 in Africa: the spread and response. Nat Med. 2020;26(7):999–1003.

7. Boum Y, Bebell LM, Bisseck A-CZ-K. Africa needs local solutions to face the COVID-19 pandemic. The Lancet. 2021;397(10281):1238–40.

8. Salyer SJ, Maeda J, Sembuche S, Kebede Y, Tshangela A, Moussif M, et al. The first and second waves of the COVID-19 pandemic in Africa: a cross-sectional study. The Lancet. 2021;397(10281):1265–75.

9. WHO. Coronavirus (COVID-19),Africa 2021. Available from: https://www.afro.who.int/health-topics/coronavirus-covid-19.

10. Gutman JR, Lucchi NW, Cantey PT, Steinhardt LC, Samuels AM, Kamb ML, et al. Malaria and Parasitic Neglected Tropical Diseases: Potential Syndemics with COVID-19? Am J Trop Med Hyg. 2020;103(2):572–7.

11. Mulenga LB, Hines JZ, Fwoloshi S, Chirwa L, Siwingwa M, Yingst S, et al. Prevalence of SARS-CoV-2 in six districts in Zambia in July, 2020: a cross-sectional cluster sample survey. The Lancet Global Health. 2021.

12. Usuf E, Roca A. Seroprevalence surveys in sub-Saharan Africa: what do they tell us? Lancet Glob Health. 2021.

13. Impouma B, Wolfe CM, Mboussou F, Farham B, Saturday T, Pervilhac C, et al. Monitoring and Evaluation of COVID-19 response in the WHO African Region: challenges and lessons learned. Epidemiol Infect. 2021:1–14.

14. Müller O. Malaria in Africa: challenges for control and elimination in the 21st century: Peter Lang Frankfurt; 2011.

15. Global Burden of Disease, Viz Hub [Internet]. University of Washington. 2021 [cited 30.04.2021]. Available from: https://vizhub.healthdata.org/gbd-compare/.

16. Amimo F, Lambert B, Magit A. What does the COVID-19 pandemic mean for HIV, tuberculosis, and malaria control? Trop Med Health. 2020;48:32.

17. WHO. World malaria report 2020: 20 years of global progress and challenges.2020 11.05.2020. Available from: https://www.who.int/teams/global-malaria-programme/reports/world-malaria-report-2020.

18. Munn Z, Peters MDJ, Stern C, Tufanaru C, McArthur A, Aromataris E. Systematic review or scoping review? Guidance for authors when choosing between a systematic or scoping review approach. BMC Medical Research Methodology. 2018;18(1):143.

19. Tricco AC, Lillie E, Zarin W, O’Brien KK, Colquhoun H, Levac D, et al. PRISMA Extension for Scoping Reviews (PRISMA-ScR): Checklist and Explanation. Ann Intern Med. 2018;169(7):467–73.

20. Schardt C, Adams MB, Owens T, Keitz S, Fontelo P. Utilization of the PICO framework to improve searching PubMed for clinical questions. BMC Med Inform Decis Mak. 2007;7:16.

21. Weiss DJ, Bertozzi-Villa A, Rumisha SF, Amratia P, Arambepola R, Battle KE, et al. Indirect effects of the COVID-19 pandemic on malaria intervention coverage, morbidity, and mortality in Africa: a geospatial modelling analysis. Lancet Infect Dis. 2020.

22. WHO. The potential impact of health service disruptions on the burden of malaria: a modelling analysis for countries in sub-Saharan Africa. 2020.

23. Sherrard-Smith E, Hogan AB, Hamlet A, Watson OJ, Whittaker C, Winskill P, et al. The potential public health consequences of COVID-19 on malaria in Africa. Nat Med. 2020;26(9):1411–6.

24. Bell D, Hansen KS. Relative burdens of the COVID-19, malaria, tuberculosis and HIV/AIDS epidemics in sub-Saharan Africa. medRxiv. 2021:2021.03.27.21254483.

25. Ansumana R, Sankoh O, Zumla A. Effects of disruption from COVID-19 on antimalarial strategies. Nat Med. 2020;26(9):1334–6.

26. Chanda-Kapata P, Kapata N, Zumla A. COVID-19 and malaria: A symptom screening challenge for malaria endemic countries. Int J Infect Dis. 2020;94:151–3.

27. Kusotera T, Nhengu TG. Coronavirus-19 and malaria: The great mimics. Afr J Prim Health Care Fam Med. 2020;12(1):e1–e3.

28. Ajayi IO, Ajumobi OO, Falade C. Malaria and COVID-19: commonalities, intersections and implications for sustaining malaria control. The Pan African medical journal. 2020;37(Suppl 1):1.

29. Buonsenso D, Iodice F, Cinicola B, Raffaelli F, Sowa S, Ricciardi W. Management of malaria in children under 5-years-old during COVID-19 pandemic in Sierra Leone: a lesson learned? medRxiv. 2020:2020.11.04.20225714.

30. WHO. Tailoring malaria interventions in the COVID-19 response. Global Malaria Programme [Internet]. 2020 05.04.2021. Available from: https://www.who.int/publications/m/item/tailoring-malaria-interventions-in-the-covid-19-response.

31. Steketee RW, Choi M, Linn A, Florey L, Murphy M, Panjabi R. World Malaria Day 2021: Commemorating 15 Years of Contribution by the United States President’s Malaria Initiative. The American Journal of Tropical Medicine and Hygiene. 2021.

32. The Global Fund. THE IMPACT OF COVID-19 ON HIV, TB AND MALARIA SERVICES AND SYSTEMS FOR HEALTH: A SNAPSHOT FROM 502 HEALTH FACILITIES ACROSS AFRICA AND ASIA 2021. Available from: https://www.theglobalfund.org/en/updates/other-updates/2021-04-13-the-impact-of-covid-19-on-hiv-tb-and-malaria-services-and-systems-for-health/.

33. Brooke B, Raman J, Frean J, Rundle K, Maartens F, Misiani E, et al. Implementing malaria control in South Africa, Eswatini and southern Mozambique during the COVID-19 pandemic. SAMJ: South African Medical Journal. 2020;110(11):1072–6.

34. Anjorin AA, Abioye AI, Asowata OE, Soipe A, Kazeem MI, Adesanya IO, et al. Comorbidities and the COVID-19 Pandemic Dynamics in Africa. Trop Med Int Health. 2020.

35. Di Gennaro F, Marotta C, Locantore P, Pizzol D, Putoto G. Malaria and COVID-19: Common and Different Findings. Trop Med Infect Dis. 2020;5(3).

36. Hussein MIH, Albashir AAD, Elawad Oama, Homeida A. Malaria and COVID-19: unmasking their ties. Malaria journal. 2020;19(1):457.

37. Beshir KB, Grignard L, Hajissa K, Mohammed A, Nurhussein AM, Ishengoma DS, et al. Emergence of Undetectable Malaria Parasites: A Threat under the Radar amid the COVID-19 Pandemic? Am J Trop Med Hyg. 2020;103(2):558–60.

38. Abdul-Mumin A, Abubakari A, Agbozo F, Abdul-Karim A, Nuertey BD, Mumuni K, et al. Field evaluation of specificity and sensitivity of a standard SARS-CoV-2 antigen rapid diagnostic test: A prospective study at a teaching hospital in Northern Ghana. medRxiv. 2021:2021.06.03.21258300.

39. Cao R, Hu H, Li Y, Wang X, Xu M, Liu J, et al. Anti-SARS-CoV-2 Potential of Artemisinins In Vitro. ACS Infect Dis. 2020;6(9):2524–31.

40. Cortegiani A, Ingoglia G, Ippolito M, Giarratano A, Einav S. A systematic review on the efficacy and safety of chloroquine for the treatment of COVID-19. J Crit Care. 2020;57:279–83.

41. Gendrot M, Duflot I, Boxberger M, Delandre O, Jardot P, Le Bideau M, et al. Antimalarial artemisinin-based combination therapies (ACT) and COVID-19 in Africa: In vitro inhibition of SARS-CoV-2 replication by mefloquine-artesunate. Int J Infect Dis. 2020;99:437–40.

42. Vincent MJ, Bergeron E, Benjannet S, Erickson BR, Rollin PE, Ksiazek TG, et al. Chloroquine is a potent inhibitor of SARS coronavirus infection and spread. Virol J. 2005;2:69-.

43. Boulware DR, Pullen MF, Bangdiwala AS, Pastick KA, Lofgren SM, Okafor EC, et al. A Randomized Trial of Hydroxychloroquine as Postexposure Prophylaxis for Covid-19. New England Journal of Medicine. 2020;383(6):517–25.

44. Cavalcanti AB, Zampieri FG, Rosa RG, Azevedo LCP, Veiga VC, Avezum A, et al. Hydroxychloroquine with or without Azithromycin in Mild-to-Moderate Covid-19. New England Journal of Medicine. 2020;383(21):2041–52.

45. Rakedzon S, Neuberger A, Domb AJ, Petersiel N, Schwartz E. From hydroxychloroquine to ivermectin: what are the anti-viral properties of anti-parasitic drugs to combat SARS-CoV-2? J Travel Med. 2021;28(2).

46. Tang W, Cao Z, Han M, Wang Z, Chen J, Sun W, et al. Hydroxychloroquine in patients with mainly mild to moderate coronavirus disease 2019: open label, randomised controlled trial. BMJ. 2020;369:m1849.

47. Kangbai JB, Sao Babawo L, Kaitibi D, Sandi AA, George AM, Sahr F. Re-reading ACT, BCG, and Low COVID-19 in Africa. SN comprehensive clinical medicine. 2021;3(1):11–5.

48. Aborode AT, David KB, Uwishema O, Nathaniel AL, Imisioluwa JO, Onigbinde SB, et al. Fighting COVID-19 at the Expense of Malaria in Africa: The Consequences and Policy Options. Am J Trop Med Hyg. 2020.

49. Nghochuzie NN, Olwal CO, Udoakang AJ, Amenga-Etego LN, Amambua-Ngwa A. Pausing the Fight Against Malaria to Combat the COVID-19 Pandemic in Africa: Is the Future of Malaria Bleak? Front Microbiol. 2020;11:1476.

50. Mbunge E, Millham R, Sibiya MN, Takavarasha S. Impact of COVID-19 on Malaria Elimination: Juxtaposing Indoor Residual Spraying and Mobile Phones in Buhera Rural District, Zimbabwe. 2021.

51. Rogerson SJ, Beeson JG, Laman M, Poespoprodjo JR, William T, Simpson JA, et al. Identifying and combating the impacts of COVID-19 on malaria. BMC Med. 2020;18(1):239.

52. Emmanuel Awucha N, Chinelo Janefrances O, Chima Meshach A, Chiamaka Henrietta J, Ibilolia Daniel A, Esther Chidiebere N. Impact of the COVID-19 Pandemic on Consumers’ Access to Essential Medicines in Nigeria. Am J Trop Med Hyg. 2020;103(4):1630–4.

53. Amimo F, Lambert B, Magit A, Hashizume M. The potential impact of the COVID-19 pandemic on HIV, tuberculosis, and malaria control in Africa: a systematic review of modelling studies and population surveys. 2020.

54. Aïkpon R, Affoukou C, Hounpkatin B, Eclou DD, Cyaka Y, Egwu E, et al. Digitalized mass distribution campaign of insecticide-treated nets (ITNs) in the particular context of Covid-19 pandemic in Benin: challenges and lessons learned. Malar J. 2020;19(1):431.

55. Shi B, Zheng J, Xia S, Lin S, Wang X, Liu Y, et al. Accessing the syndemic of COVID-19 and malaria intervention in Africa. 2020.

56. Baral S, Rao A, Twahirwa Rwema JO, Lyons C, Cevik M, Kågesten AE, et al. Competing Health Risks Associated with the COVID-19 Pandemic and Response: A Scoping Review. medRxiv. 2021:2021.01.07.21249419.

57. Guerra CA, Tresor Donfack O, Motobe Vaz L, Mba Nlang JA, Nze Nchama LO, Mba Eyono JN, et al. Malaria vector control in sub-Saharan Africa in the time of COVID-19: no room for complacency. BMJ Glob Health. 2020;5(9).

58. The Alliance for Malaria Prevention. 2020 Annual Report 2020. Available from: https://allianceformalariaprevention.com/wp-content/uploads/2021/03/FINAL-AMP-Annual-Report-2020.pdf.

59. Newby G, Mpanju-Shumbusho W, Feachem RGA. Global health security requires endemic disease eradication. The Lancet. 2021;397(10280):1163–5.

60. RBM Partnership to end malaria. RBM Partnership to End Malaria position on the next Global Fund strategy2020. Available from: https://endmalaria.org/sites/default/files/RBM%20Position%20Statement%20on%20Global%20Fund%27s%20Strategy.pdf.

61. Bell D, Hansen KS, Kiragga AN, Kambugu A, Kissa J, Mbonye AK. Predicting the Impact of COVID-19 and the Potential Impact of the Public Health Response on Disease Burden in Uganda. Am J Trop Med Hyg. 2020;103(3):1191–7.

62. Burt J, Ouma J, Amone A, Aol L, Sekikubo M, Nakimuli A, et al. Indirect Effects of COVID-19 on Maternal, Neonatal, Child, Sexual and Reproductive Health Services in Kampala, Uganda. medRxiv. 2021:2021.04.23.21255940.

63. Hategeka C, Carter SE, Chenge FM, Katanga EN, Lurton G, Mayaka SM-N, et al. Impact of the COVID-19 pandemic and response on the utilisation of health services during the first wave in Kinshasa, the Democratic Republic of the Congo. medRxiv. 2021:2021.04.08.21255096.

64. Gavi S, Tapera O, Mberikunashe J, Kanyangarara M. Malaria incidence and mortality in Zimbabwe during the COVID-19 pandemic: analysis of routine surveillance data. Malar J. 2021;20(1):233.

65. Chasaya M PM, Ngomah MA. An Update on Malaria trends in Zambia (2019 to 2020); A descriptive study. Health Press Zambia Bull. 2020:13–8.

66. Müller O, Razum O, Jahn A. Effects of non-pharmaceutical interventions against COVID-19 on the incidence of other diseases. The Lancet Regional Health – Europe. 20216.

67. Abbas K, Mogasale V. Disruptions to childhood immunisation due to the COVID-19 pandemic. The Lancet.

68. Zawawi A, Alghanmi M, Alsaady I, Gattan H, Zakai H, Couper K. The impact of COVID-19 pandemic on malaria elimination. Parasite Epidemiology and Control. 2020;11:e00187.

69. Wang J, Xu C, Wong YK, He Y, Adegnika AA, Kremsner PG, et al. Preparedness is essential for malaria-endemic regions during the COVID-19 pandemic. Lancet. 2020;395(10230):1094–6.

70. Velavan TP, Meyer CG, Esen M, Kremsner PG, Ntoumi F, Consortium P-I-NC. COVID-19 and syndemic challenges in & lsquo;Battling the Big Three & rsquo;: HIV, TB and malaria. International Journal of Infectious Diseases. 2021;106:29–32.

71. Nkeshimana M, Nsanzimana S. Malaria or COVID-19? A case report highlighting a diagnostic challenge in Africa. 2021.

72. Achan Jane SA, Wanzira Humphrey, Kyagulanyi Tonny, Nuwa Anthony, Magumba Godfrey, Kusasira Stephen, Sewanyana Isaac, Tetteh Kevin, Drakeley Chris, Nakwagala Fredrick, Aanyu Helen, Opigo Jimmy, Hamade Prudence, Marasciulo Madeleine, Baterana Byarugaba, Tibenderana, James,. Impact of Current Malaria Infection and Previous Malaria Exposure on the Clinical Profiles and Outcome of COVID-19 in a High Malaria Transmission Setting: A Prospective Cohort Study. The Lancet pre-prints SSRN. 2021.

73. C Coban Kji, T Kawai, H Hemmi, S Sato, S Uematsu, Frosch AE, John CC. Immunomodulation in Plasmodium falciparum malaria: experiments in nature and their conflicting implications for potential therapeutic agents. Expert review of anti-infective therapy. 2005;201.

74. Thompson MG, Breiman RF, Hamel MJ, Desai M, Emukule G, Khagayi S, et al. Influenza and Malaria Coinfection Among Young Children in Western Kenya, 2009–2011. The Journal of Infectious Diseases. 2012;206(11):1674–84.

75. Edwards CL, Zhang V, Werder RB, Best SE, Sebina I, James KR, et al. Coinfection with Blood-Stage Plasmodium Promotes Systemic Type I Interferon Production during Pneumovirus Infection but Impairs Inflammation and Viral Control in the Lung. Clinical and Vaccine Immunology. 2015;22(5):477–83.

76. Hogan AB, Jewell BL, Sherrard-Smith E, Vesga JF, Watson OJ, Whittaker C, et al. Potential impact of the COVID-19 pandemic on HIV, tuberculosis, and malaria in low-income and middle-income countries: a modelling study. Lancet Glob Health. 2020;8(9):e1132–e41.

77. Figueroa JP, Bottazzi ME, Hotez P, Batista C, Ergonul O, Gilbert S, et al. Urgent needs of low-income and middle-income countries for COVID-19 vaccines and therapeutics. The Lancet. 2021;397(10274):562–4.

78. The Lancet Infectious D. The rocky road to universal COVID-19 vaccination. The Lancet Infectious Diseases. 2021;21(6):743.

79. Rahi M, Das P, Sharma A. COVID-19 Mitigation Steps Provide a Blueprint for Malaria Control and Elimination. Am J Trop Med Hyg. 2020;103(1):28–30.

80. Buonsenso D, Cinicola B, Kallon MN, Iodice F. Child Healthcare and Immunizations in Sub-Saharan Africa During the COVID-19 Pandemic. Front Pediatr. 2020;8:517.

81. Coker M, Folayan MO, Michelow IC, Oladokun RE, Torbunde N, Sam-Agudu NA. Things must not fall apart: the ripple effects of the COVID-19 pandemic on children in sub-Saharan Africa. Pediatr Res. 2020.

82. Diongue K, Diallo MA. COVID-19 during malaria transmission season in Africa and why we should be prepared: An example from Senegal. African Journal of Laboratory Medicine. 2020;9(1):3.

83. Inzaule SC, Tessema SK, Kebede Y, Ouma AEO, Nkengasong JN. Genomic-informed pathogen surveillance in Africa: opportunities and challenges. The Lancet Infectious Diseases. 2021.

84. Makanjuola RO, Ishaleku D, Taylor-Robinson A. COVID-19 and malaria in sub-saharan Africa: Holistic diagnostic approaches may promote effective clinical case management. Microbes and Infectious Diseases. 2020;1(3):100–6.

85. Mvumbi DM. Mass intake of hydroxychloroquine or chloroquine in the present context of the Covid-19 outbreak: Possible consequences in endemic malaria settings. Med Hypotheses. 2020;143:109912.

86. P.M.I. US President’s malaria initiative. U.S. President’s Malaria Initiative Burkina Faso Malaria Operational Plan FY 20202020. Available from: www.pmi.gov.

87. Rosenthal PJ, Breman JG, Djimde AA, John CC, Kamya MR, Leke RGF, et al. COVID-19: Shining the Light on Africa. Am J Trop Med Hyg. 2020;102(6):1145–8.

88. The Global Fund. MITIGATING THE IMPACT OF COVID-19 ON COUNTRIES AFFECTED BY HIV, TUBERCULOSIS AND MALARIA2020. Available from: https://www.theglobalfund.org/media/9819/covid19_mitigatingimpact_report_en.pdf.

